# Quantifying Human–AI Workflow in Abdominal Ultrasound: A Prospective Randomised Crossover Study

**DOI:** 10.64898/2026.08.17.26360254

**Authors:** Natalie Hsiao, Mehrnaz Clifford, Shang-Zhe Lin, Swapna Premasiri, Jacqueline Roots, Heather Allen, Aaron P. Robertson, Khalid Moafa, Jane Wardle, Christopher Edwards

## Abstract

**Objective:** To evaluate the effect of vendor-integrated AI-assisted abdominal ultrasound software on operational efficiency and sonographer workload compared with manual scanning.

**Methods:** In this prospective randomised crossover study (January to February 2026), 32 healthy adults each underwent two upper abdominal examinations, one manual and one using vendor-integrated AI software (AI Abdomen Release 3.5; ACUSON Sequoia), in randomised order by two experienced sonographers; each participant was scanned once by each sonographer. Scan time, hand–console interaction (keystrokes, hand travel, hover, jerk) from a custom depth-camera hand-tracking system, and operator modifications to AI outputs were recorded. Workload was assessed after each scan with the weighted NASA Task Load Index (NASA-TLX). Analysis used linear mixed-effects models.

**Results:** AI-assisted scanning reduced scan time (52.4 s, approximately 9%; 95% CI 23.7 to 81.2; *P* = 0.001), keystrokes (55, approximately 28%; *P* < 0.001) and hand travel (4.57 m, approximately 39%; *P* < 0.001), although the time saving was concentrated in one sonographer. Weighted NASA-TLX did not differ between conditions (−3.9 points; 95% CI −9.3 to 1.5; *P* = 0.17), but subscale analyses showed reductions in mental demand (−6.3; *P* = 0.03) and effort (−7.0; *P* = 0.04), with no compensating increases. Sonographers modified 48 of 184 AI-generated values.

**Conclusion:** AI assistance improved operational efficiency and reduced self-reported mental demand and effort, with no compensating increase on other subscales. Gains arose under a controlled, abbreviated protocol in healthy volunteers and varied between operators, and are better read as a reshaping of operator work than its removal.

## INTRODUCTION

In diagnostic ultrasound, the image does not exist until a skilled operator creates it, manipulating the probe, interpreting in real time, and working the console to optimise, acquire, label, and measure ^1^. That dependence on operator skill is also its constraint. As applications widen and demand continues to rise ^2^ the manual dexterity and cognitive stamina the work requires cannot be manufactured at the rate the workforce needs ^3^.

Work-related musculoskeletal disorders (WRMSDs) are highly prevalent among sonographers ^4^. These injuries have historically been attributed to mechanical factors, including repetitive movements, static postures, and sustained console interaction, but evidence increasingly implicates cognitive and psychosocial load as well ^5–7^. The demand to identify the correct view and obtain measurements manually is not only physical but also cognitive, requiring sustained perceptual processing and decision-making throughout the examination ^1^.

Manufacturers have responded with artificial intelligence (AI) tools that support image optimisation during scanning ^8–10^. Some of these systems recognise standard anatomical views and generate preliminary measurements for key organs, functioning as real-time assistants while the clinician retains control of probe manipulation, optimisation, and interpretation. Most imaging AI interprets a stored study, such as a CT or radiograph, whereas AI-assisted ultrasound acts during acquisition, alongside the clinician, under the same clinical uncertainty that defines the examination ^11^.

Early independent evidence from cardiac applications suggests such systems can reduce scan time and console interaction ^12^. However, the impact of AI-assisted ultrasound extends beyond efficiency alone. Acquisition, interpretation, and decision-making occur concurrently under human control, making AI-assisted ultrasound a genuine instance of human–AI collaboration ^9^. Its effectiveness depends not on algorithmic performance alone but on how it redistributes operator effort: trading the manual demands of acquisition for the work of checking, and sometimes correcting, its outputs. It also depends on whether operators trust those outputs enough to act on them ^13^.

Organisational, cultural, medicolegal, and user-acceptance factors all shape how technology is implemented ^14–16^; here we set these aside to focus on the real-time interaction between operator and AI under controlled conditions. Our aim was to examine how AI-assisted abdominal ultrasound changes scanning workflow, with particular attention to operational efficiency and operator workload. We hypothesised that AI assistance would improve workflow efficiency, reduce console-side physical interaction, and shift cognitive demand towards image optimisation, verification, and quality assurance. We also characterised the AI-generated measurement outputs and how they may shape the operator’s experience of scanning.

## MATERIALS AND METHODS

### Study design and ethical approval

This prospective randomised crossover simulation study evaluated the workflow effects of an AI-assisted abdominal ultrasound system under controlled scanning conditions. A crossover design was chosen so that each participant served as their own control, removing between-participant variation in body habitus and sonographic access, which are dominant sources of variance in scan duration and operator effort. Reporting followed the CONSORT 2025 statement ^17^, and its extension for randomised crossover trials ^18^, with the DECIDE-AI checklist ^19^ used to guide reporting of the AI system and its use, adapted for this simulated setting. The study protocol, approved by the institutional Human Research Ethics Committee on 15 December 2025 prior to data collection, pre-specified scan duration and weighted NASA-TLX as the primary outcomes. Keystrokes, hand-travel distance, hover time, and jerk were secondary operational workflow outcomes; all remaining analyses, including the efficiency– workload association, operator-modification burden, subscale-level NASA-TLX analyses, and sensitivity analyses, were exploratory. The study was not prospectively registered in a clinical trials registry; participants were not assigned to a health-related intervention to evaluate health outcomes, and the outcomes evaluated were operator workload and workflow. No multiplicity adjustment was applied. Examinations were designated non-diagnostic in the approved protocol, and no clinical decisions were made on AI output; adverse events were therefore not systematically assessed. A referral pathway for formal diagnostic imaging was in place should an unexpected finding be observed during an examination. Participant data were de-identified using sequentially coded identifiers (USAI-01 to USAI-32), with no direct identifiers retained in the analysis dataset.

### Participants

Two accredited diagnostic medical sonographers with 11 and 25 years of adult abdominal ultrasound experience performed the examinations. Neither had used the specific AI-assisted system under evaluation before the study. Each completed a single standardised vendor-delivered training session on the system before data collection, with no further clinical exposure to it.

Healthy adult volunteers aged 18 years or older were recruited by convenience sampling from the university community. Individuals with known significant abdominal pathology, prior major abdominal surgery likely to alter anatomy, or contraindications to diagnostic ultrasound were excluded. Participants were asked to fast four hours before scanning; one attended non-fasted and was scanned as planned. Scanning was performed in an ultrasound laboratory between January and February 2026. No a priori power calculation was performed. Post hoc, 32 within-participant pairs provided 80% power (α = 0.05, two-sided) to detect dz = 0.51, approximately 7.9 NASA-TLX points; the study was therefore underpowered for smaller workload effects.

Sex was recorded as participant-reported sex assigned at birth; gender identity was not collected. Sex was treated as a descriptive participant characteristic rather than an outcome variable, and the study was neither designed nor powered to examine sex differences in scanning performance.

### Scanning workflow and AI system

Each participant was randomly allocated to one of four sequences (n = 8 per sequence) using a computer-generated random sequence in Microsoft Excel, with both condition order (AI-first vs manual-first) and sonographer order independently randomised and balanced (**Figure 1; Supplementary Table S1**). Each participant was scanned once by each sonographer, with AI-assisted and manual examinations performed by different sonographers. The two examinations were performed consecutively within a single session, separated by a five-minute pause allowing the participant a short rest and the sonographers to change over. Across the cohort, each sonographer performed 16 AI-assisted and 16 manual scans. The allocation sequence was generated by a research assistant and held on a dedicated laptop accessible only to them; sonographers were not informed of a participant’s assigned sequence until the time of each scan. Sonographers could not be blinded, because the AI-assisted workflow is visually distinct on the console, and the data analyst was present during data collection and was therefore also unblinded. The AI system (AI Abdomen Release 3.5 [VB30]) was implemented as vendor-integrated software on the ultrasound platform (ACUSON Sequoia, Siemens Medical Solutions USA, Inc., Mountain View, CA, USA) using a 5C1 curvilinear transducer and the standard vendor abdominal preset ^20^. No model training, tuning, or parameter optimisation was performed. The algorithm class, the population on which the algorithm was trained, and its preclinical performance are not publicly disclosed by the manufacturer. The software version, ultrasound system, transducer, and vendor abdominal preset were unchanged for the duration of data collection; no vendor software update or hardware servicing occurred during the study. During scanning, the system generated anatomical labels and preliminary measurements; sonographers retained full control and could accept, modify, or reject these outputs.

**Figure 1.**
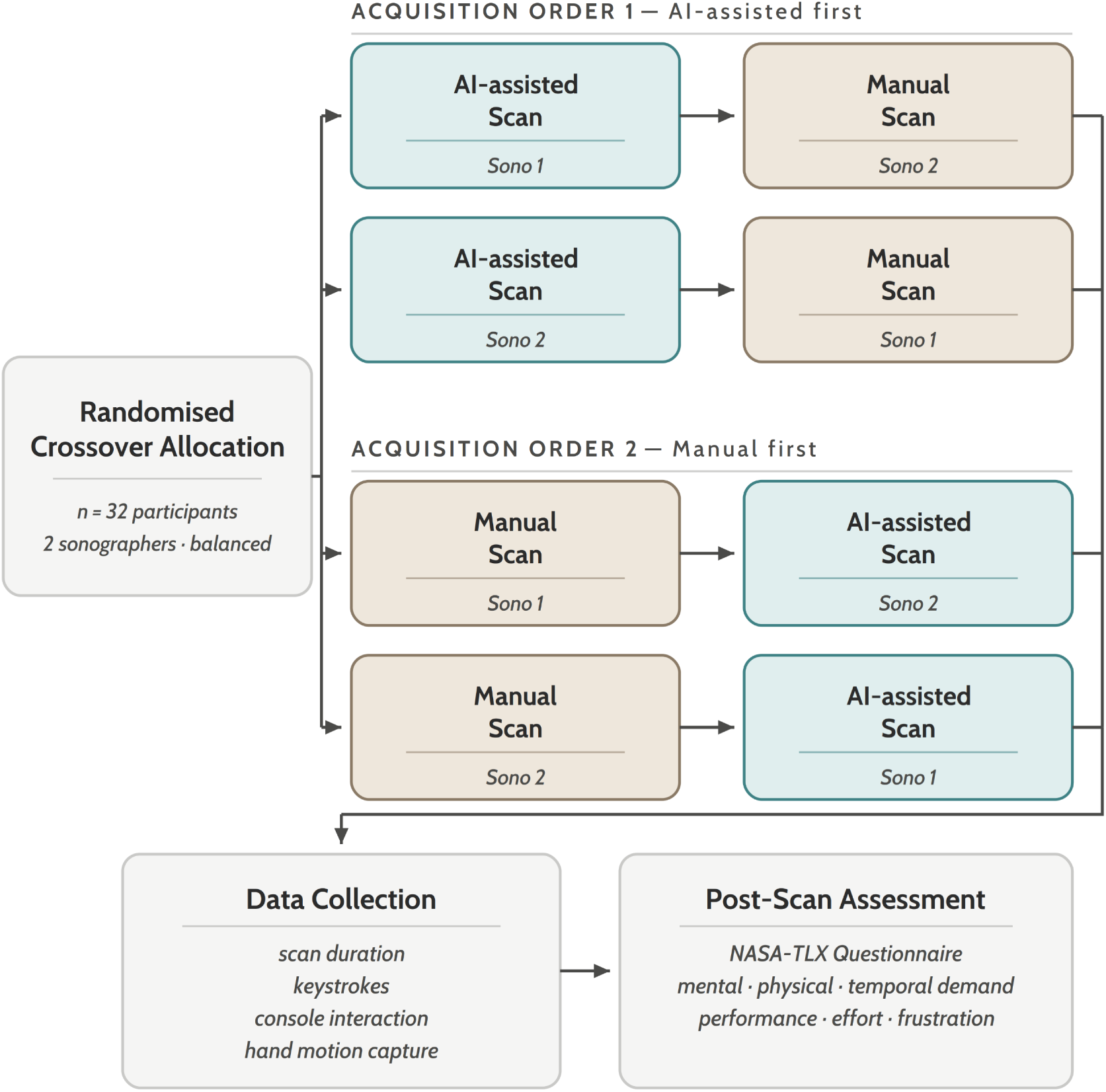
Study workflow. Each participant underwent two upper abdominal ultrasound examinations — one AI-assisted and one manual, each performed by a different sonographer — and was randomly allocated to one of four sequences (n = 8 per sequence), with both condition order and sonographer order randomised and balanced. Each sonographer performed 16 AI-assisted and 16 manual scans across the cohort (Supplementary Table S1). Workflow metrics (scan duration, keystrokes, console interaction, and hand motion) were recorded during scanning, and perceived workload was assessed after each scan using the NASA-TLX.

The application operates on the live B-mode stream during freehand scanning. It recognises the anatomical view being imaged, applies an anatomical label, and positions measurement callipers on the corresponding structure to generate a preliminary biometric value. When the label and callipers were rendered, the sonographer may accept, adjust or reject them before the image is stored.

Imaging depth, gain and time-gain compensation were adjusted by the operator during scanning as required for body habitus; transmit frequency, focal configuration, tissue harmonic imaging and acoustic output remained at preset defaults for both conditions.

Both scan conditions followed a standard upper abdominal ultrasound protocol comprising longitudinal and transverse views of the pancreas, aorta, inferior vena cava, liver, gallbladder, common bile duct, kidneys, and spleen, with routine biometric measurements restricted to those supported by the AI system (**Figure 2; Supplementary Table S2**). Doppler assessment was excluded because Doppler functions were not available within the AI-assisted workflow.

**Figure 2.**
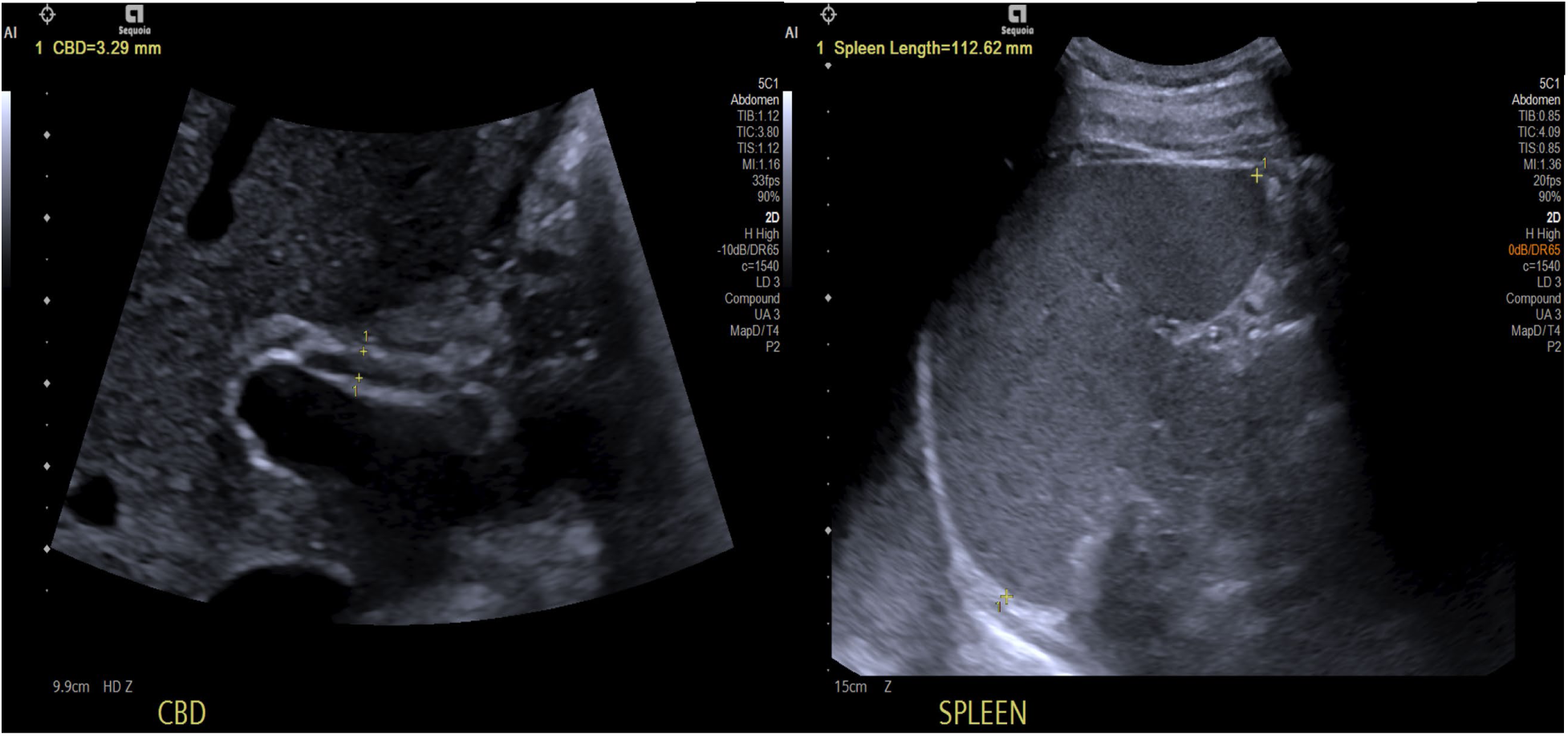
Ultrasound images of the common bile duct (CBD) and spleen displaying AI-generated labels and measurements.

### Operator workload and interaction metrics

Perceived workload was assessed after each scan using the NASA Task Load Index (NASA-TLX), a multidimensional instrument comprising six domains: mental demand, physical demand, temporal demand, performance, effort, and frustration ^21^. Ratings were completed immediately after each examination, followed by the pairwise weighting procedure, with weighted scores calculated by standard NASA-TLX methodology ^21^. Each subscale was also rescaled to 0–100 and retained for subscale-level analysis alongside the weighted composite.

Hand–keyboard interaction was quantified with a custom Python hand-tracking system (HTS)^22^ using a fixed depth camera above the console, yielding four per-scan metrics: cumulative hand travel distance, hover time in the keyboard zone, average movement jerk, and scan duration (first probe contact to acquisition of the final required image; the same recording window was used for all metrics). Keystrokes were counted in real time by an independent observer. Acquisition, preprocessing, metric formulas, the heatmap method, and a face-validity check are detailed in **Supplementary Appendix S1**, and the system is illustrated in **Supplementary Figure S1**.

The system used an Intel RealSense D435 depth camera mounted in a fixed position above the console, acquiring synchronised colour and depth streams at 30 frames per second at a resolution of 848 x 480 pixels. Sonographers wore dark blue nitrile gloves; glove pixels were isolated by thresholding in hue-saturation-value colour space, and a channel transformation (green attenuated by 50%, red set to zero) shifted the segmented pixels towards the skin-tone profile expected by the MediaPipe Hand Landmarker, which returned 21 hand landmarks per frame. Detection was confined to the central 40% of the camera field of view to exclude monitor reflections and peripheral movement. The hand centroid was taken as the mean position of the wrist and the four metacarpophalangeal landmarks and was deprojected to three dimensions using the corresponding depth value. Trajectories were despiked by rejecting inter-frame displacements exceeding 0.3 m and smoothed with a One Euro filter before cumulative travel distance, hover time and average jerk were derived by numerical differentiation. Camera position, lighting and console layout were held constant across all recordings. Full formulas, tracking yield and a face-validity check are given in **Supplementary Appendix S1**.

### Reference standard and AI output verification

Because this study evaluated AI-assisted scanning during real-time acquisition, no single external reference standard was defined. Instead, the scanning sonographer verified AI outputs in real time; in this workflow context, sonographer verification represented the clinically relevant reference process.

For each AI-supported measurement, we noted whether the AI generated a value and the action the sonographer took: no change, modification of an AI-generated value, or manual measurement after the AI generated no value. Annotation modifications, in which AI-generated anatomical labels were amended, were recorded separately and assessed when images were frozen and saved. Operator interventions per scan were the sum of these event types. Each AI-supported measurement was attempted once per examination, and modification frequency is reported against the number of examinations in which the AI generated that measurement. Formal inter- or intra-rater variability was not assessed because outputs were verified contemporaneously; measurement-agreement analyses are the subject of separate work. The AI-supported views and measurements with their confirmation requirements are summarised in **Supplementary Table S2.**

### Statistical analysis

Statistical analyses were performed in R (version 4.5.1) ^23^. Continuous outcomes are reported as mean differences with 95% confidence intervals. Counts are reported as n (%), and proportions of examinations in which an AI-generated measurement was modified are reported with 95% Wilson score confidence intervals. Differences between AI-assisted and manual scans were examined using linear mixed-effects models to account for the randomised crossover design and repeated measures within participants, with scan condition, sonographer, and scan order as fixed effects and participant as a random intercept. These models were applied to efficiency, hand-interaction, and workload outcomes (weighted NASA-TLX and each of the six subscale scores, modelled separately. Sensitivity analyses additionally fitted a condition-by-sonographer interaction term (**Supplementary Table S3**), and included a study-sequence covariate to assess any trend across the study period; neither altered the main scan-duration or weighted NASA-TLX estimates. Standardised coefficients shown in Figure 3 are the condition effect from the same model fitted to each outcome scaled to unit variance, expressed as the AI-assisted minus manual difference in standard deviation units.

**Figure 3.**
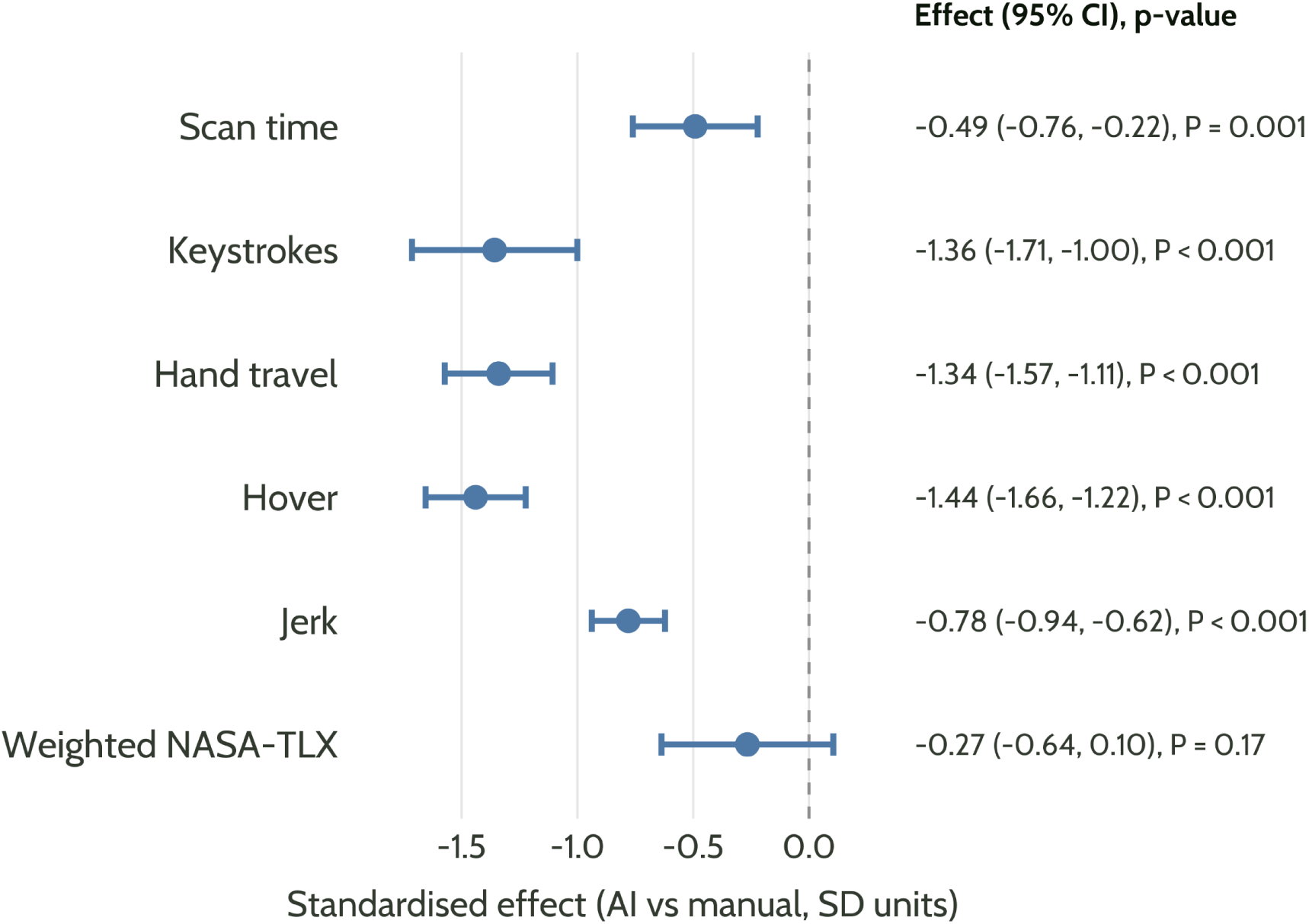
Forest plot showing standardised model coefficients for primary and secondary operational workflow outcomes (AI-assisted vs manual scanning) from linear mixed-effects models. Estimates are expressed in standard deviation units to allow comparison across outcomes measured on different scales. Bars indicate 95% confidence intervals.

In exploratory analyses, two further models examined the relationship between scanning workflow and workload. First, weighted NASA-TLX was modelled as a function of scan duration in a linear mixed-effects model (scan condition and sonographer as fixed effects; participant as a random intercept) to examine within-participant covariation. Second, weighted NASA-TLX was modelled as a function of the total number of operator interventions, restricted to AI-assisted scans and adjusted for scan duration and sonographer. Sensitivity to individual participants was assessed for the primary outcome models and for both regressions using leave-one-participant-out re-fits; the range of the focal coefficient across all 32 fits is reported. Linear mixed-effects models were fitted using the lme4 package ^24^, with degrees of freedom and p-values estimated using lmerTest ^25^. As a robustness check, paired non-parametric Wilcoxon signed-rank tests were performed on the primary and secondary operational workflow outcomes (**Supplementary Table S4**). No missing data occurred for the primary outcomes, and all tests were two-sided, with a nominal significance level of α = 0.05.

## RESULTS

### Participants and allocation

A total of 32 participants underwent 64 abdominal ultrasound examinations (manual n = 32, AI-assisted n = 32; Table 1). All protocol-required views and measurements were obtained in every manual examination (192/192). In AI-assisted examinations, 189 of 192 required measurements were obtained; one was omitted by the operator and two were not recorded for reasons not documented. Efficiency gains therefore reflected faster completion of the same protocol rather than reduced scanning. No adverse events occurred, and no unexpected findings requiring referral were observed.

**Table 1.** Characteristics of the study population.

| *Characteristic* | *N = 32<sup>1</sup>* |
| --- | --- |
| *Sex* |  |
| *Female* | *20 (62.5%)* |
| *Male* | *12 (37.5%)* |
| *Age* | *25.3 (8.9); 18–48* |
| *BMI* | *22.6 (3.6); 16.4–31.3* |
| <sup>1</sup> *n (%)*; *Mean (SD)*; *Min–Max* |  |
| BMI = body mass index |  |

### Efficiency and console interaction

In linear mixed-effects models adjusting for sonographer, scan order, and repeated measures within participants, AI-assisted scanning was associated with shorter scan duration and reduced console-side interaction (Table 2). The primary scan-duration outcome fell by 52.4 s (95% CI 23.7–81.2; *P* = 0.001), while weighted NASA-TLX did not differ significantly between conditions (mean difference −3.9 points; 95% CI −9.3 to +1.5; *P* = 0.17). Standardised model coefficients are shown in Figure 3. The primary scan-duration finding and all four secondary operational workflow findings remained significant under paired non-parametric testing, whereas the weighted NASA-TLX comparison remained non-significant (**Supplementary Table S4**).

**Table 2.** Primary and secondary operational workflow outcomes by scan condition, with adjusted mean differences and 95% confidence intervals.

| ***Outcome*** | ***Manual mean*** | ***AI mean*** | ***Mean Change [95% CI]*** | ***% Change*** | ***P value*** |
| --- | --- | --- | --- | --- | --- |
| ***Keystrokes (n)*** | 197 | 142 | $-55.0 [-69, -41]$ | $-28\%$ | $<0.001$ |
| ***Hand travel (m)*** | 11.81 | 7.24 | $-4.57 [-5.36, -3.78]$ | $-39\%$ | $<0.001$ |
| ***Scan Time (sec)*** | 592.4 | 540 | $-52.4 [-81.2, -23.7]$ | $-9\%$ | $0.001$ |
| ***Hover (sec)*** | 64.4 | 31.1 | $-33.3 [-38.3, -28.3]$ | $-52\%$ | $<0.001$ |
| ***Jerk (m/s<sup>3</sup>)*** | 3.06 | 2.23 | $-0.83 [-0.99, -0.66]$ | $-27\%$ | $<0.001$ |
| ***Weighted total NASA- TLX*** | 45.8 | 41.9 | $-3.9 [-9.3, +1.5]$ | $-9\%$ | $0.17$ |
| *Estimates are adjusted mean differences (AI-assisted minus manual) from linear mixed-effects models with scan condition, sonographer and scan order as fixed effects and participant as a random intercept. NASA-TLX = NASA Task Load Index.* |  |  |  |  |  |

Sensitivity analyses with condition-by-sonographer interaction terms showed directionally consistent AI effects across both sonographers for all primary outcomes with a main effect, though the magnitude varied between sonographers (**Supplementary Table S3**). No single participant drove any primary estimate. In leave-one-participant-out re-fits the condition effect retained its direction in all 32 fits for every outcome. (**Supplementary Table S5**). Perceived workload declined across the study period in both conditions (AI-assisted: −0.93 NASA-TLX points per participant, *P* < 0.001; manual: −0.55, *P* = 0.04), with a weaker trend in scan time, whereas participant body mass index (BMI) was stable (*P* = 0.94); a first-half versus second-half comparison of AI-assisted scans is reported in **Supplementary Table S6**. This pattern is consistent with general familiarisation rather than condition-specific learning, and adjusting the primary workload model for study sequence did not alter the AI effect.

Spatial heatmaps (Figure 4) showed interaction concentrated around the central console in both workflows, with visually reduced density during AI-assisted scanning, consistent with the lower hand travel and hover time.

**Figure 4.**
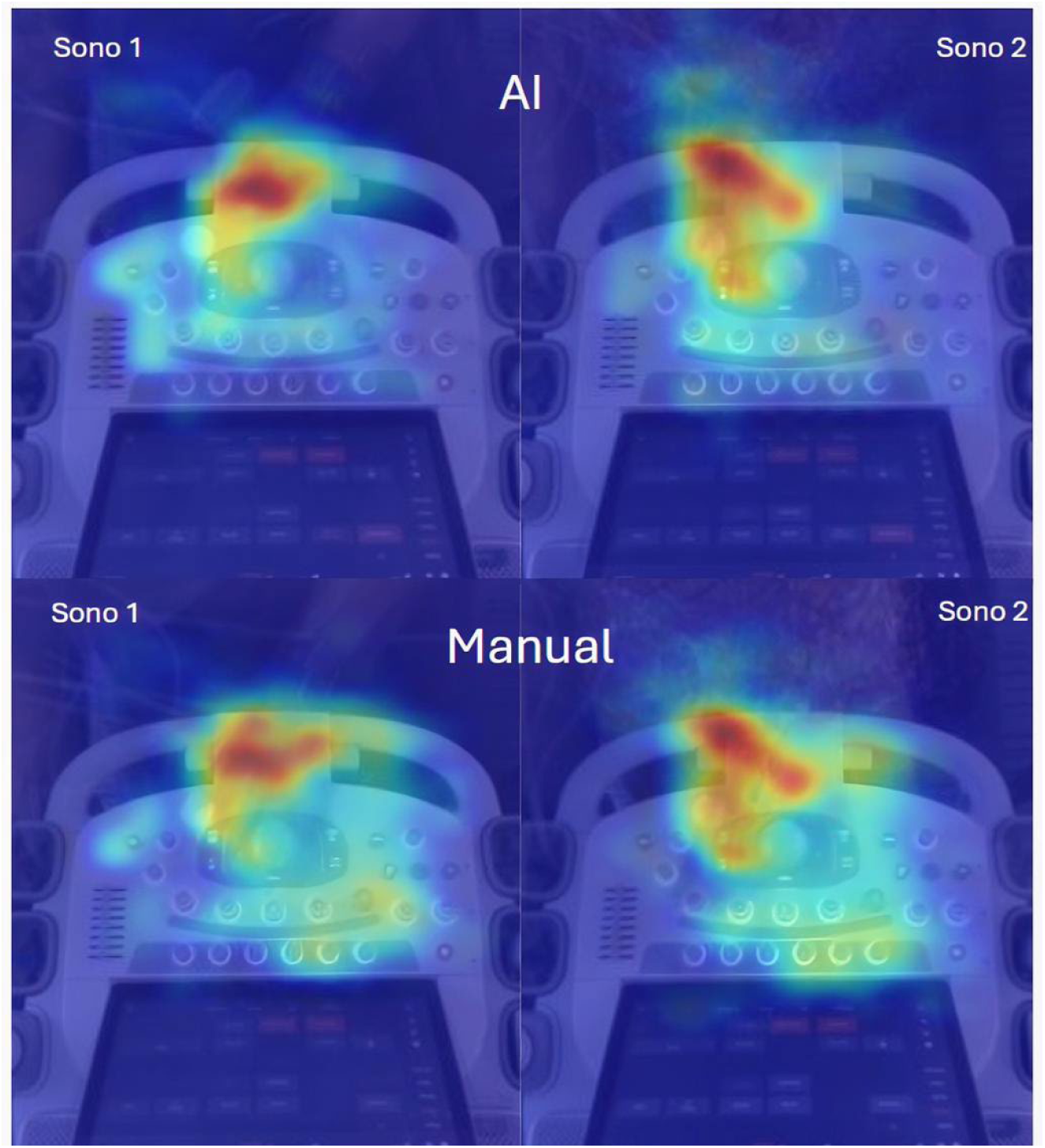
Averaged spatial heatmaps of console interaction during abdominal ultrasound scanning for two sonographers (Sono 1 and Sono 2) under AI-assisted and manual workflows. Each panel is the pixel-wise average of independently normalised per-scan heatmaps; warmer colours indicate higher relative interaction density within that panel. Comparisons reflect spatial interaction patterns.

### Perceived workload

Although the weighted composite did not differ between conditions (Table 2), subscale analyses showed significant reductions under AI assistance in mental demand (β = −6.3; *P* = 0.03) and effort (β = −7.0; *P* = 0.04), with no significant differences on the other four subscales (**Supplementary Table S7**). Scan duration was positively associated with workload within participants (β = 0.089 NASA-TLX points per second; 95% CI 0.060–0.118; *P* < 0.001; leave-one-out range 0.085–0.095), independent of condition and sonographer (**Supplementary Figure S2**).

### Operator modifications to AI output

During AI-assisted scanning, the AI generated a value for 184 of the 189 measurements obtained. In the remaining five (gallbladder wall, n = 2; common bile duct diameter, n = 2; right kidney length, n = 1) it generated no value and the operator measured manually, one gallbladder in the non-fasted participant. Sonographers modified 48 of the 184 AI-generated values and made 33 annotation modifications; only one of the 32 examinations required no operator intervention. Modification was most frequent for liver span (18/31; 58%; 95% CI, 41– 74), followed by gallbladder wall thickness (11/30; 37%; 95% CI, 22–55) and common bile duct diameter (7/29; 24%; 95% CI, 12–42), with wide and overlapping confidence intervals (Figure 5A). Both sonographers modified a similar overall proportion of AI-generated values (21/90 and 27/94), although the measurements involved differed (Figure 5B). Generation and operator action for each measurement are given in **Supplementary Table S8**.

**Figure 5.**
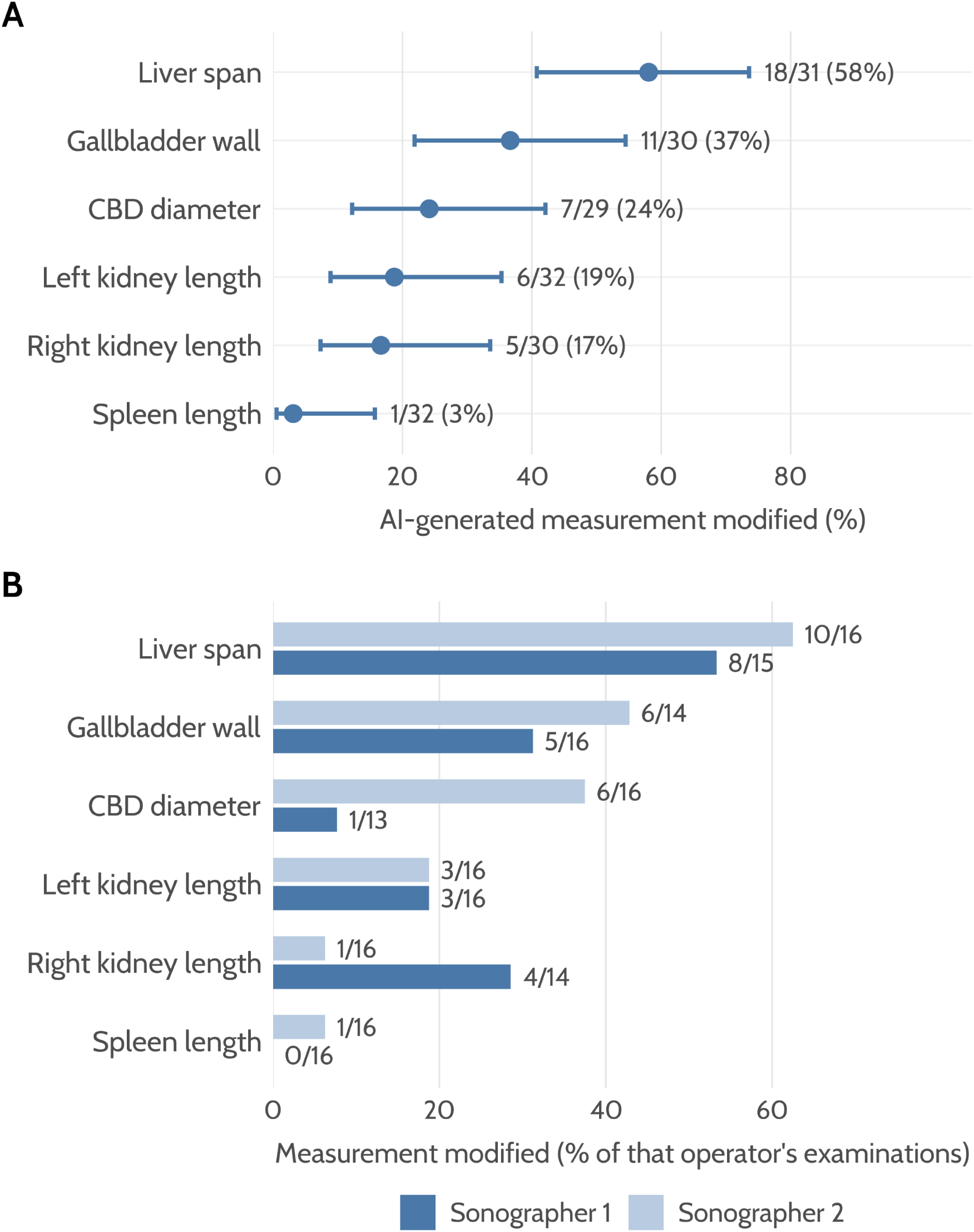
Operator modifications to AI-generated measurements during AI-assisted abdominal ultrasound scanning. (A) Proportion of examinations in which each AI-generated measurement was modified, with 95% Wilson score confidence intervals; the denominator is the number of examinations in which the AI generated that measurement. (B) The same modifications by sonographer, with 16 AI-assisted examinations performed by each. Panel B is descriptive; the study was not designed to compare operators on modification frequency.

Annotation modifications were concentrated among anatomically adjacent, sonographically confusable structures, predominantly within the epigastric vascular and ductal region (inferior vena cava, aorta, pancreas, common bile duct, and left hepatic lobe), with pancreas amended to inferior vena cava the most frequent pair (n = 4). A second, smaller group involved contralateral organs, right kidney amended to left kidney being the next most frequent pair (n = 3). The full distribution of AI-assigned and operator-amended label pairs is provided in **Supplementary Table S9**.

In an exploratory analysis restricted to AI-assisted scans, the total number of operator interventions was positively, but not significantly, associated with weighted NASA-TLX (β = 2.22 points per intervention; 95% CI, −0.35 to +4.78; *P* = 0.087; leave-one-out range, 1.79 to 2.79), adjusted for scan duration and sonographer. Scan duration remained strongly associated with workload (β = 0.10 points per second; *P* < 0.001); sonographer was not.

## DISCUSSION

In this two-operator crossover study, AI-assisted abdominal ultrasound was associated with improved operational efficiency at the group level. Comparable reductions have been reported in cardiac sonography under an AI-assisted protocol ^12^, suggesting the benefit is not confined to a single anatomical domain. Average jerk was also lower under AI assistance, consistent with smoother console-hand trajectories when fewer keystrokes and corrections are required. The time savings were modest, and whether they would alter service-level throughput is uncertain; clinic capacity also depends on unmeasured factors such as room turnover and reporting time and would need a dedicated study to establish. The composite weighted NASA-TLX did not differ significantly between conditions, and the study was underpowered to characterise it precisely; however, subscale analyses showed reductions in self-reported mental demand and effort, with no compensating increase on any subscale. In this controlled setting with experienced sonographers, AI assistance was therefore associated with reduced, rather than redistributed, perceived cognitive load.

Our workload results differ from a larger published trial of AI-assisted ultrasound acquisition. In the PROMETHEUS trial, AI assistance during the fetal anomaly scan reduced median scan duration from 19.7 to 11.4 minutes and lowered composite NASA-TLX (median 35.2 versus 46.5) ^26^. Our composite estimate was lower by only 3.9 points and did not reach significance, although mental demand and effort fell at the subscale level. Several differences plausibly account for the gap. The PROMETHEUS tool automated the freeze, save and measure cycle across 13 standard planes and four biometrics, removing a larger share of manual work than the annotation and measurement support evaluated here. Its image capture also ran silently during acquisition, with human review of candidate frames deferred until the scan was complete, whereas the system evaluated here displays labels and callipers on the live image and asks the operator to accept, adjust or reject them while scanning. Verification demand remained concurrent with acquisition in our study. PROMETHEUS additionally randomised sonographers to method, so its workload contrast is between operators, while ours is within operator and rests on two sonographers. Cognitive load theory would predict the pattern we observed if the AI removed extraneous, operational demand^27^ and returned part of it as concurrent verification, which the automation literature anticipates ^28,29^.

PROMETHEUS also offers the closest available evidence on image quality under acquisition-assist AI. Image quality of the images selected by the AI was initially rated below manually acquired images. When an improved selection model was applied retrospectively to the same recorded examinations, quality matched manual acquisition for many planes ³⁷. Two implications follow for real-time systems. Output quality depends on the curation and presentation step as much as on the underlying model, and operator review can only correct what the interface offers. In the PROMETHEUS trial, the shortfall was visible only because the trial performed an independent quality assessment, which is the check our design did not include. The trial’s authors propose running quality models in real time for this reason ^26^.

The modifications recorded during AI-assisted scanning show a clinically meaningful pattern. They concentrated on sonographically confusable structures, indicating that mislabelling occurred often enough to warrant ongoing vigilance. Two patterns are distinguishable. Most errors involved structures lying adjacent within the same imaging plane, predominantly the epigastric vessels and ducts. A smaller group involved contralateral organs of near-identical sonographic appearance. The AI generated no value in five instances. Each recorded modification and each detected non-generation represents an AI failure the operator caught. How often incorrect outputs went uncorrected cannot be established from this design. Less experienced sonographers, less able to recognise such errors, would carry a greater risk of accepting incorrect output ^30^.

For patients, operator-level effects would plausibly translate into shorter waiting times, more examinations completed in a list, and more time for the studies that are difficult to complete. These gains are indirect, however, and depend on the time saving persisting in clinically representative examinations, on image quality and measurement accuracy held under AI assistance, and on the saved time being returned to the schedule rather than absorbed into the working day.

### Limitations

Examinations were performed on healthy, low-BMI volunteers using an abbreviated protocol that excluded Doppler, so efficiency gains may not transfer to older or heavier patients, significant bowel gas, or pathology, though this population suited an initial task-level evaluation. Formal image quality and measurement accuracy were not assessed; a quality-for-speed trade-off cannot be excluded and is the focus of separate work. Both sonographers were highly experienced, and the workload pattern may differ for less experienced users. The hand-tracking metrics reflect keyboard-hand interaction only; scanning-arm posture and probe-hand force were not measured, and the findings should not be read as evidence of reduced musculoskeletal injury risk. Sonographers could not be blinded because the AI workflow is visually distinct on the console.

Operator exposure to the system before the study was limited to a single standardised vendor training session and one practice scan, and within-study analyses showed improvement in perceived workload and movement smoothness across AI scans, consistent with operators continuing to adapt. The within-participant crossover design protects the AI-versus-manual contrast from this longitudinal trend, although the absolute magnitude of the cognitive workload effects should be interpreted accordingly. With only two sonographers, condition and sonographer were partially confounded at the participant level. We addressed this through balanced allocation, sonographer and scan order as fixed effects, and condition-by-sonographer interaction models as sensitivity analyses. The AI effect was directionally consistent across sonographers for all primary outcomes but varied in magnitude, with the scan time and weighted NASA-TLX effects concentrated in one sonographer. As this study evaluated a single on-cart AI solution, some effects may not extend to other platforms.

## CONCLUSION

Vendor-integrated AI reduced scan duration, keystrokes and hand travel during abdominal ultrasound, and lowered self-reported mental demand and effort with no compensating increase on any other subscale. In this controlled setting with experienced operators, real-time AI assistance reduced the operational work of scanning, and no shift into supervisory effort was detectable. The correction burden concentrated on sonographically confusable structures, which indicates where interface design and selective deployment of the more reliable functions would pay off. Whether these gains reach patients depends on their persistence in clinically representative examinations, on evidence that image quality and measurement accuracy hold, and on how any saved time is used within the service. Evaluation should extend to less experienced operators and to patients with pathology, and should assess AI output alongside the interface that presents it.

## ETHICS APPROVAL AND CONSENT TO PARTICIPATE

The study was approved by the Queensland University of Technology Human Research Ethics Committee on 15 December 2025 (approval number 10419), before data collection, and was conducted in accordance with the Declaration of Helsinki. All participants provided written informed consent, including consent for de-identified data arising from the study to be published. Patients and members of the public were not involved in the design, conduct, or reporting of this study.

## ACKNOWLEDGEMENTS

The authors thank the volunteers who participated in this study and acknowledge the technical support provided by the Quantitative Ultrasound Imaging Laboratory at Queensland University of Technology.

## DATA AVAILABILITY STATEMENT

The de-identified dataset and the R analysis code supporting the findings of this study are deposited with the QUT Research Data Finder and are available under mediated access through the request workflow described in the record https://doi.org/10.25912/RDF_1786424222552. The custom hand-tracking software (HTS) used to derive the interaction metrics is publicly available at https://github.com/LolMaple/HTS. All analyses reported in this article are repeatable using the archived data.

## SUPPLEMENTARY MATERIAL

Supplementary Appendix S1 (hand-tracking system: acquisition, preprocessing, metric definitions, heatmap method and face-validity check), Supplementary Tables S1–S9 and Supplementary Figures S1–S2 are provided in a separate file uploaded with this preprint.

## REFERENCES

1 Nicholls, D., Sweet, L. & Hyett, J. Psychomotor skills in medical ultrasound imaging: an analysis of the core skill set. Journal of Ultrasound in Medicine 33, 1349–1352 (2014). 10.7863/ultra.33.8.1349

2 Clevert, D. A., Beyer, G., Niess, H. & Schlenker, B. Ultrasound-New Techniques Are Extending the Applications. Dtsch Arztebl Int 120, 41–47 (2023). 10.3238/arztebl.m2022.0380

3 Edwards, C., Tunny, R., Allen, H., Bowles, D., Farley, A., O’Hara, S., Wardle, J. & Reddan, T. Sonographer training pathways-a discussion paper on curriculum design and implementation. International Journal of Work-Integrated Learning 25, 321–336 (2024).

4 Zangiabadi, Z., Makki, F., Marzban, H., Salehinejad, F., Sahebi, A. & Tahernejad, S. Musculoskeletal disorders among sonographers: a systematic review and meta-analysis. BMC Health Services Research 24, 1233 (2024). 10.1186/s12913-024-11666-w

5 Xie, Y., Coombes, B. K., Thomas, L. & Johnston, V. Musculoskeletal Pain and Disability in Sonographers: More Than an Ergonomic Issue. Journal of the American Society of Echocardiography 33, 1526–1527 (2020). 10.1016/j.echo.2020.07.005

6 Gibbs, V. & Edwards, H. An investigation of sonographers unaffected by work-related musculoskeletal disorders. Ultrasound 20, 149–154 (2012). 10.1258/ult.2012.012014

7 Fukumura, Y. E., Sommerich, C. M., Evans, K. D. & Roll, S. C. Work-Related Musculoskeletal Disorders and Associated Work Systems Factors: Are There Differences Between Sonography Practice Areas? Journal of Diagnostic Medical Sonography 40, 4–18 (2023). 10.1177/87564793231205612

8 Edwards, C., Chamunyonga, C., Searle, B. & Reddan, T. The application of artificial intelligence in the sonography profession: Professional and educational considerations. Ultrasound 30, 273–282 (2022). 10.1177/1742271X211072473

9 Day, T. G., Matthew, J., Budd, S., Hajnal, J. V., Simpson, J. M., Razavi, R. & Kainz, B. Sonographer interaction with artificial intelligence: collaboration or conflict? Ultrasound in Obstetrics and Gynecology 62, 167–174 (2023). 10.1002/uog.26238

10 Cui, X. W., Goudie, A., Blaivas, M., Chai, Y. J., Chammas, M. C., Dong, Y., Stewart, J., Jiang, T. A., Liang, P., Sehgal, C. M., Wu, X. L., Hsieh, P. C., Adrian, S. & Dietrich, C. F. WFUMB Commentary Paper on Artificial intelligence in Medical Ultrasound Imaging. Ultrasound in Medicine and Biology 51, 428–438 (2025). 10.1016/j.ultrasmedbio.2024.10.016

11 Erickson, B. J., Korfiatis, P., Akkus, Z. & Kline, T. L. Machine Learning for Medical Imaging. Radiographics 37, 505–515 (2017). 10.1148/rg.2017160130

12 Hollitt, K. J., Milanese, S., Joseph, M. & Perry, R. Can automation and artificial intelligence reduce echocardiography scan time and ultrasound system interaction? Echo Res Pract 12, 11 (2025). 10.1186/s44156-025-00077-0

13 Vaccaro, M., Almaatouq, A. & Malone, T. When combinations of humans and AI are useful: A systematic review and meta-analysis. Nat Hum Behav 8, 2293–2303 (2024). 10.1038/s41562-024-02024-1

14 Hua, D., Petrina, N., Young, N., Cho, J. G. & Poon, S. K. Understanding the factors influencing acceptability of AI in medical imaging domains among healthcare professionals: A scoping review. Artificial Intelligence in Medicine 147, 102698 (2024). 10.1016/j.artmed.2023.102698

15 Naicker, S., Schmidt, P., Shar, B., Tariq, A., Earnshaw, A. & McPhail, S. Implementing an Artificial Intelligence Decision Support System in Radiology: Prospective Qualitative Evaluation Study Using the Nonadoption Abandonment Scale-Up, Spread, and Sustainability (NASSS) Framework. Journal of Medical Internet Research 28, e80342 (2026). 10.2196/80342

16 Ross, J., Hammouche, S., Chen, Y., Rockall, A. G. & Royal College of Radiologists, A. I. W. G. Beyond regulatory compliance: evaluating radiology artificial intelligence applications in deployment. Clinical Radiology 79, 338–345 (2024). 10.1016/j.crad.2024.01.026

17 Hopewell, S., Chan, A. W., Collins, G. S., Hrobjartsson, A., Moher, D., Schulz, K. F., Tunn, R., Aggarwal, R., Berkwits, M., Berlin, J. A., Bhandari, N., Butcher, N. J., Campbell, M. K., Chidebe, R. C. W., Elbourne, D., Farmer, A., Fergusson, D. A., Golub, R. M., Goodman, S. N., Hoffmann, T. C., Ioannidis, J. P. A., Kahan, B. C., Knowles, R. L., Lamb, S. E., Lewis, S., Loder, E., Offringa, M., Ravaud, P., Richards, D. P., Rockhold, F. W., Schriger, D. L., Siegried, N. L., Staniszewska, S., Taylor, R. S., Thabane, L., Torgerson, D., Vohra, S., White, I. R. & Boutron, I. CONSORT 2025 statement: Updated guideline for reporting randomised trials. PLoS Medicine 22, e1004587 (2025). 10.1371/journal.pmed.1004587

18 Dwan, K., Li, T., Altman, D. G. & Elbourne, D. CONSORT 2010 statement: extension to randomised crossover trials. BMJ 366, l4378 (2019). 10.1136/bmj.l4378

19 Vasey, B., Nagendran, M., Campbell, B., Clifton, D. A., Collins, G. S., Denaxas, S., Denniston, A. K., Faes, L., Geerts, B., Ibrahim, M., Liu, X., Mateen, B. A., Mathur, P., McCradden, M. D., Morgan, L., Ordish, J., Rogers, C., Saria, S., Ting, D. S. W., Watkinson, P., Weber, W., Wheatstone, P., McCulloch, P. & group, D.-A. e. Reporting guideline for the early stage clinical evaluation of decision support systems driven by artificial intelligence: DECIDE-AI. BMJ 377, e070904 (2022). 10.1136/bmj-2022-070904

20 Siemens Healthineers. *Siemens Healthineers introduces industry-first AI Abdomen as part of ACUSON Sequoia 3.5 ultrasound*, <https://www.siemens-healthineers.com/en-ie/press-room/press-features/acuson-sequoia-3.5> (2024).

21 Hart, S. G. & Staveland, L. E. in Human mental workload. Advances in psychology, 52. 139–183 (North-Holland, 1988).

22 Lin, S.-Z. *HTS: Hand Tracking System*, <https://github.com/LolMaple/HTS> (2026).

23 R: A Language and Environment for Statistical Computing (R Foundation for Statistical Computing, Vienna, Austria, 2025).

24 Bates, D., Mächler, M., Bolker, B. & Walker, S. Fitting Linear Mixed-Effects Models Using lme4. Journal of Statistical Software 67 (2015). 10.18637/jss.v067.i01

25 Kuznetsova, A., Brockhoff, P. B. & Christensen, R. H. B. lmerTest Package: Tests in Linear Mixed Effects Models. Journal of Statistical Software 82, 1–26 (2017). 10.18637/jss.v082.i13

26 Day, T. G., Matthew, J., Budd, S. F., Farruggia, A., Venturini, L., Wright, R., Jamshidi, B., To, M., Ling, H., Lai, J., Tan, M. Y., Brown, M., Guy, G., Casagrandi, D., Arechvo, A., Syngelaki, A., Lloyd, D., Zidere, V., Vigneswaran, T., Miller, O., Akolekar, R., Nanda, S., Nicolaides, K., Kainz, B., Simpson, J. M., Hajnal, J. V. & Razavi, R. AI to Assist in the Fetal Anomaly Ultrasound Scan: A Randomized Controlled Trial. Nejm Ai 2, AIoa2400747 (2025). 10.1056/AIoa2400747

27 Sweller, J., Van Merrienboer, J. J. & Paas, F. G. Cognitive architecture and instructional design. Educational Psychology Review 10, 251–296 (1998). 10.1023/A:1022193728205

28 Bainbridge, L. Ironies of automation. Automatica 19, 775–779 (1983). 10.1016/0005-1098(83)90046-8

29 Parasuraman, R., Sheridan, T. B. & Wickens, C. D. A model for types and levels of human interaction with automation. IEEE Trans Syst Man Cybern A Syst Hum 30, 286–297 (2000). 10.1109/3468.844354

30 Dratsch, T., Chen, X., Rezazade Mehrizi, M., Kloeckner, R., Mahringer-Kunz, A., Pusken, M., Baessler, B., Sauer, S., Maintz, D. & Pinto Dos Santos, D. Automation Bias in Mammography: The Impact of Artificial Intelligence BI-RADS Suggestions on Reader Performance. Radiology 307, e222176 (2023). 10.1148/radiol.222176

