## Supplementary Methods for "Quantifying Human–AI Workflow in Abdominal Ultrasound: A Prospective Randomised Crossover Study"

### Appendix S1 — Hand-tracking acquisition and preprocessing

#### System overview

Hand–keyboard interaction was quantified using a custom Hand-Tracking System (HTS) implemented in Python [1]. The system used an Intel RealSense D435 depth-sensing camera mounted in a fixed position above the ultrasound console, with the camera position held constant across all recordings to enable valid spatial heatmap comparisons. Video and depth streams were acquired at 30 frames per second (dt ≈ 0.033 s) at a resolution of 848 × 480 pixels, with neutral lighting maintained throughout to ensure consistent colour-based hand segmentation. Recordings were saved as MJPEG-compressed AVI files (OpenCV VideoWriter, MJPG fourcc, 30 fps). MJPEG applies per-frame lossy compression with no inter-frame dependencies, preserving per-frame image quality suitable for re-analysis.

#### Hand detection and segmentation

Sonographers wore dark blue nitrile gloves to provide high colour contrast against the ultrasound console and patient skin, enabling reliable glove segmentation by thresholding in the hue–saturation–value (HSV) colour space. To improve detection reliability, a colour-channel transformation was then applied to the masked image: the green channel was attenuated by 50% and the red channel was set to zero (OpenCV BGR channels 1 and 2). This shifted the cyan-blue glove pixels towards a skin-like profile that the MediaPipe model was trained to detect. The original colour image was retained for all display, recording, and heatmap outputs.

The colour-transformed, glove-segmented inference image passed to MediaPipe was restricted to the central 40% of the camera field of view, with the left and right 30% blanked, confining detection to the operator–console interaction zone and excluding monitor reflections and peripheral movement. Within this region, MediaPipe Hand Landmarker (Google) [2] identified 21 hand landmarks per frame. The model (VIDEO running mode) was configured with minimum hand-detection, presence, and tracking confidence of 0.30, below the default of 0.50, increasing detection sensitivity in the colour-transformed, glove-masked images at the cost of occasional false detections.

The hand centroid was computed as the mean pixel position of five MediaPipe landmarks: the wrist and the four metacarpophalangeal (MCP) joints (landmark indices 0, 5, 9, 13, 17), approximating the geometric centre of the palm. Three-dimensional coordinates (X, Y, Z) were obtained by deprojecting the centroid pixel location using the corresponding depth value from the RealSense depth stream, where X is horizontal (right positive), Y vertical (down positive), and Z depth (forward positive, relative to the camera centre). Video recordings and spatial heatmaps were cropped separately to the central 50% (left and right 25%) for display and comparison only; this crop did not affect the detection region used in the analysis.

#### Preprocessing pipeline

Raw 3D centroid trajectories were processed in the following sequence:

1. Jump rejection. Movements exceeding 0.3 m between consecutive frames were treated as sensor noise or transient tracking errors and excluded from metric derivation.

2. Adaptive smoothing. A One Euro Filter [3] was applied to the 3D coordinates to reduce frame-to-frame jitter while preserving rapid intentional movements (parameters: $\beta = 0.1$, $f_{min} = 0.01$ Hz, $f_d = 1.0$ Hz).

3. Metric derivation. Spatiotemporal metrics were computed on the filtered centroid trajectory.

**Metric definitions and discrete-time formulas**

For each scan, four metrics were derived from the filtered centroid trajectory:

Scan duration (s): Total elapsed time of the active tracking session, accumulated as the sum of inter-frame intervals (dt) during active tracking only. Tracking was started and stopped manually by the operator, from first probe contact with the participant's skin to acquisition of the final required image.

Cumulative hand travel distance (m): Sum of three-dimensional Euclidean distances between consecutive valid centroid positions:

$$D = \Sigma \|P_t - P_{t-1}\|$$

Hover time (s): Cumulative duration during which the hand was both (a) close to the camera/console (depth $Z < 0.75$ m) and (b) actively moving (instantaneous speed $S \geq 0.05$ m/s), where $S = \|P_t - P_{t-1}\| / dt$. Hover time approximated active console interaction time, distinct from static rest periods. When both conditions were met in a given frame, dt was added to hover time.

Average jerk ($m/s^3$): Magnitude of the third numerical derivative of position, averaged across all valid frames:

$$V_t = (P_t - P_{t-1}) / dt$$

$$A_t = (V_t - V_{t-1}) / dt$$

$$J_t = (A_t - A_{t-1}) / dt$$

$$\text{Average jerk} = (1/N) \cdot \Sigma \|J_t\|$$

Lower values indicate smoother, more controlled motion; higher values indicate more abrupt or erratic changes in acceleration.

Keystrokes were recorded manually for each examination as a separate measure of operator–console interaction, independent of the hand-tracking pipeline.

**Heatmap generation**

Spatial console-interaction heatmaps were generated for each scan by projecting the X–Y coordinates of the hand centroid onto the camera frame plane at each frame and accumulating frequencies into a two-dimensional histogram. Bin counts were log-transformed (log(1 + count)) to compress the dynamic range, so that both high-dwell hotspots and sparse movement paths remained visible. The histogram was then smoothed with a Gaussian blur, colour-mapped, and overlaid on a reference console image. Each heatmap was normalised using per-session min–max scaling. Heatmap colour, therefore, represents the relative log-density of centroid positions within a single scan, supporting interpretation of spatial interaction patterns. Depth was not incorporated, so centroid positions at different distances from the camera are not distinguished in the projected plane.

To generate the per-condition visualisations in Figure 4, the independently normalised heatmaps were aligned to the common camera reference frame and averaged pixel-wise across all examinations within each condition × sonographer cell. Because the contributing heatmaps were normalised within session rather than to a shared global maximum, the four panels are not on a common intensity scale.

They should be read as a comparison of the spatial distribution of console interaction, not of absolute interaction density between panels.

#### Missing-frame handling and tracking yield

Frames excluded by the jump-rejection rule or where MediaPipe failed to detect a hand were omitted from metric computation. Per-scan tracking yield was not separately quantified per condition; the deterministic processing pipeline applied identical exclusion and smoothing rules regardless of scan condition, so any condition-related differences in derived metrics reflect the underlying movement patterns rather than systematic differences in tracking performance between conditions.

#### Face-validity check

As a face-validity check, the relationship between cumulative hand travel distance (measured by the hand-tracking system) and the independently recorded keystroke count was examined across all 64 examinations. The two measures were significantly positively correlated (Pearson $r = 0.42$, 95% CI 0.19 to 0.60, $P < .001$; Spearman $\rho = 0.68$, $P < .001$), consistent with both reflecting the intensity of operator–console interaction during scanning. The higher rank correlation (Spearman) than linear correlation (Pearson) suggests a monotonic but non-strictly-linear relationship between the two measures.

#### Reliability and validation

Formal validation of the hand-tracking system against an external reference standard (e.g., marker-based optical motion capture) was beyond the scope of this study. The system was developed as a research tool to characterise relative changes in console-side hand interaction under different scanning conditions, rather than to provide absolute biomechanical measurements. Test–retest and inter-observer reliability were not formally assessed, as recordings were processed by a single automated pipeline with deterministic outputs given fixed video input. The face-validity check above (hand travel vs keystroke count) supports the system's coherence as an index of console interaction; formal validation against a gold-standard reference is a direction for further methodological work.

### Familiarisation protocol

Prior to study commencement, both sonographers completed a brief familiarisation session with the AI-assisted system using a non-study volunteer (one practice scan per sonographer on the same volunteer), to ensure baseline competence with the system's interface, prompts, and correction workflow. No formal vendor training was undertaken. The absence of an extended familiarisation period is acknowledged as a limitation in the main text.

### Sensitivity analyses

Two exploratory regressions reported in the main text (the scan-level scan time–workload model and the AI-only edit-burden model) were assessed for sensitivity to individual participants using leave-one-participant-out (LOPO) re-fitting. The scan-level scan time–workload model was re-fitted 32 times, each time omitting one participant (two scans). For the AI-only edit-burden model, the regression was re-fitted 32 times, each time omitting one participant's single AI scan. The range of the focal coefficient across all 32 LOPO fits was used to characterise stability.

For the scan-level scan time–workload model, the time coefficient ranged from 0.085 to 0.095 (SD 0.003) across the 32 LOPO fits, well within the full-data 95% confidence interval (0.060–0.118); no single participant influenced the result. For the AI-only edit-burden model, the intervention coefficient ranged from 1.79 to 2.79 (SD 0.24) across the 32 LOPO fits and remained positive in every fit; the point estimate is stable, and the borderline non-significance reflects the smaller AI-only subset (n = 32) rather than fragility of the underlying association.

#### Early versus late AI scans

To address whether AI-assisted scanning improved across the study period as the sonographers developed familiarity with the system, AI scans were divided into the first 16 (“early”) and the last 16 (“late”) in order of acquisition and compared on each primary outcome using independent-samples t-tests (Supplementary Table S6). Operational efficiency outcomes (scan time, keystrokes, hand travel, hover) were stable between early and late AI scans (all $P$ >.10). However, both perceived workload (mean Late − Early difference: −15.9 NASA-TLX points; P = 0.002) and movement jerk (−1.20 m/s³; $P$ < 0.001) were significantly lower in late AI scans, consistent with the sonographers acclimatising to the system and requiring less corrective hand movement over the course of the study, rather than gaining raw efficiency. The within-participant crossover design ensures that the AI-vs-manual contrast for each participant is not directly confounded by this longitudinal trend, but the absolute magnitude of the AI cognitive-workload effects should be interpreted considering this within-study learning.

### Supplementary Tables

#### Table S1 — Sonographer–condition allocation

Each participant was randomly allocated to one of four sequences (n = 8 per sequence) using a computer-generated random sequence, with both condition order (AI-first vs manual-first) and sonographer order randomised and balanced. The full per-participant allocation is shown below.

| Participant | Scan 1 condition | Scan 1 sonographer | Scan 2 condition | Scan 2 sonographer |
|---|---|---|---|---|
| USAI-01 | AI | Sono 1 | Manual | Sono 2 |
| USAI-02 | AI | Sono 2 | Manual | Sono 1 |
| USAI-03 | Manual | Sono 1 | AI | Sono 2 |
| USAI-04 | Manual | Sono 2 | AI | Sono 1 |
| USAI-05 | AI | Sono 1 | Manual | Sono 2 |
| USAI-06 | AI | Sono 2 | Manual | Sono 1 |
| USAI-07 | Manual | Sono 1 | AI | Sono 2 |
| USAI-08 | Manual | Sono 2 | AI | Sono 1 |
| USAI-09 | AI | Sono 1 | Manual | Sono 2 |
| USAI-10 | AI | Sono 2 | Manual | Sono 1 |
| USAI-11 | Manual | Sono 1 | AI | Sono 2 |
| USAI-12 | Manual | Sono 2 | AI | Sono 1 |

| Participant | Scan 1 condition | Scan 1 sonographer | Scan 2 condition | Scan 2 sonographer |
|---|---|---|---|---|
| USAI-13 | AI | Sono 1 | Manual | Sono 2 |
| USAI-14 | AI | Sono 2 | Manual | Sono 1 |
| USAI-15 | Manual | Sono 1 | AI | Sono 2 |
| USAI-16 | Manual | Sono 2 | AI | Sono 1 |
| USAI-17 | AI | Sono 1 | Manual | Sono 2 |
| USAI-18 | AI | Sono 2 | Manual | Sono 1 |
| USAI-19 | Manual | Sono 1 | AI | Sono 2 |
| USAI-20 | Manual | Sono 2 | AI | Sono 1 |
| USAI-21 | AI | Sono 1 | Manual | Sono 2 |
| USAI-22 | AI | Sono 2 | Manual | Sono 1 |
| USAI-23 | Manual | Sono 1 | AI | Sono 2 |
| USAI-24 | Manual | Sono 2 | AI | Sono 1 |
| USAI-25 | AI | Sono 1 | Manual | Sono 2 |
| USAI-26 | AI | Sono 2 | Manual | Sono 1 |
| USAI-27 | Manual | Sono 1 | AI | Sono 2 |
| USAI-28 | Manual | Sono 2 | AI | Sono 1 |
| USAI-29 | AI | Sono 1 | Manual | Sono 2 |
| USAI-30 | AI | Sono 2 | Manual | Sono 1 |
| USAI-31 | Manual | Sono 1 | AI | Sono 2 |
| USAI-32 | Manual | Sono 2 | AI | Sono 1 |

Allocation was balanced: Sonographer 1 performed 16 AI-assisted and 16 manual scans; Sonographer 2 performed 16 AI-assisted and 16 manual scans.

### Table S2 — AI-supported anatomical views and measurements

The AI-assisted system (AI Abdomen Release 3.5 [VB30], ACUSON Sequoia; Siemens Healthineers) recognised the views and generated the measurements listed below. Output was applied automatically, with no accept step; the sonographer could edit or delete it, and nothing entered the study record unless the image was stored. No prompt was given when a measurement failed to generate, so a non-generation appeared only as an absent value the sonographer had to notice and measure manually. This table contextualises the edit-burden findings in the main text (Figure 5, Supplementary Table S9).

| **Anatomical structure** | **View(s) recognised** | **Measurement(s) generated** |
|---|---|---|
| Liver | Left lobe & Right lobe, Transverse, Longitudinal | Liver span (right lobe craniocaudal) |
| Gallbladder | Longitudinal, Transverse | Gallbladder wall thickness |
| Common bile duct | Longitudinal | CBD diameter |
| Right kidney | Longitudinal, Transverse | Renal length (longitudinal) |
| Left kidney | Longitudinal, Transverse | Renal length (longitudinal) |
| Aorta | Longitudinal | None |
| Inferior vena cava | Longitudinal | None |
| Pancreas | Transverse | None |
| Spleen | Longitudinal | Splenic length |

### Table S3 — Condition × Sonographer sensitivity analysis

To assess whether the AI association varied by sonographer, linear mixed-effects models for the primary and secondary operational workflow outcomes were re-fitted with a condition × sonographer interaction term. Main effects are reported for each sonographer separately, alongside the interaction term and likelihood-ratio test p-value.

| **Outcome** | **Main-effect AI [95% CI], *P*** | **Sono 1: AI effect [95% CI], *P*** | **Sono 2: AI effect [95% CI], *P*** | **Interaction (Sono 2 vs Sono 1) [95% CI]** | **LRT *P*** |
|---|---|---|---|---|---|
| Keystrokes (n) | −55.0 [−69.2, −40.8], <.001 | −66.6 [−87.2, −45.9], <.001 | −43.4 [−64.1, −22.8], <.001 | +23.1 [−4.7, +51.0] | .102 |
| Scan time (s) | −52.4 [−80.6, −24.2], .001 | −106.6 [−178.1, −35.0], 0.005 | +1.8 [−69.8, +73.3], .961 | +108.3 [−17.9, +234.6] | .090 |
| Hand travel (m) | −4.57 [−5.35, −3.79], <.001 | −5.94 [−7.64, −4.24], <.001 | −3.20 [−4.90, −1.50], <.001 | +2.74 [−0.16, +5.63] | .064 |
| Hover (s) | −33.30 [−38.21, −28.39], <.001 | −44.40 [−54.60, −34.20], <.001 | −22.20 [−32.40, −12.00], <.001 | +22.20 [+4.98, +39.42] | .013 |

| Outcome | Main-effect AI [95% CI], *P* | Sono 1: AI effect [95% CI], *P* | Sono 2: AI effect [95% CI], *P* | Interaction (Sono 2 vs Sono 1) [95% CI] | LRT *P* |
|---|---|---|---|---|---|
| Jerk ($m/s^3$) | −0.83 [−0.99, −0.66], <.001 | −0.64 [−1.33, +0.06], .071 | −1.02 [−1.71, −0.33], .005 | −0.38 [−1.68, +0.91] | .550 |
| Weighted NASA-TLX | −3.89 [−9.31, +1.53], .170 | −10.90 [−20.89, −0.91], .037 | +3.11 [−6.88, +13.11], .543 | +14.01 [−2.37, +30.39] | .097 |

### Table S4 — Non-parametric robustness check

As a robustness check against the distributional assumptions of the primary linear mixed-effects models, paired non-parametric Wilcoxon signed-rank tests were performed on each primary outcome. Results agreed directionally and substantively with the mixed-effects analyses for all six outcomes, confirming that the primary findings are not driven by distributional assumptions.

| Outcome | Wilcoxon V | Wilcoxon *P* | Mixed-effects *P* (primary) | Agreement |
|---|---|---|---|---|
| Scan time (s) | 115 | .005 | .001 | ✓ |
| Keystrokes (n) | 0 | <.001 | <.001 | ✓ |
| Hand travel (m) | 4 | <.001 | <.001 | ✓ |
| Hover (s) | 0 | <.001 | <.001 | ✓ |
| Jerk ($m/s^3$) | 14 | <.001 | <.001 | ✓ |
| Weighted NASA-TLX | 189 | .17 | .17 | ✓ |

#### Table S5 — Leave-one-participant-out sensitivity

Each primary outcome model was re-fitted 32 times, omitting one participant per fit, to test whether any single participant accounted for the condition effect. The table reports the full-sample estimate, the range of the condition coefficient across the 32 fits, the standard deviation of those estimates, and whether the effect retained its direction in every fit. The direction held for all six outcomes, and no estimate depended on the inclusion of any individual participant.

| Outcome | Full-sample estimate | Leave-one-out range | SD across fits | Direction retained |
|---|---|---|---|---|
| Scan time (s) | −52.406 | −58.450 to −46.134 | 2.767 | Yes |
| Keystrokes (n) | −55.000 | −57.437 to −48.679 | 1.501 | Yes |
| Hand travel (m) | −4.569 | −4.732 to −4.388 | 0.076 | Yes |
| Hover time (s) | −33.300 | −34.714 to −32.209 | 0.482 | Yes |
| Jerk ($m/s^3$) | −0.827 | −0.864 to −0.788 | 0.016 | Yes |
| Weighted NASA-TLX | −3.891 | −5.135 to −2.912 | 0.522 | Yes |

*Estimates are the AI-assisted minus manual condition coefficient from the primary linear mixed-effects model for each outcome.*

#### Table S6 — Early versus late AI scans

Independent-samples t-tests comparing the first 16 versus the last 16 AI scans (in order of acquisition) on each primary outcome. Operational efficiency outcomes were stable across the study period; perceived workload and movement jerk improved significantly in late AI scans, consistent with the sonographers acclimatising to the system rather than gaining raw efficiency.

| Outcome | Early AI (first 16) | Late AI (last 16) | Late − Early difference | *P* |
|---|---|---|---|---|
| Scan time (s) | 564.9 | 515.0 | −49.9 | .12 |
| Keystrokes (n) | 143.6 | 139.4 | −4.3 | .58 |
| Hand travel (m) | 7.09 | 7.39 | +0.30 | .71 |
| Hover (s) | 29.9 | 32.3 | +2.4 | .53 |
| Jerk ($m/s^3$) | 2.83 | 1.63 | −1.20 | <.001 |
| Weighted NASA-TLX | 49.8 | 33.9 | −15.9 | .001 |

**Table S7 — NASA-TLX subscale results**

In addition to the weighted composite NASA-TLX, each of the six raw subscales was rescaled to 0–100 and modelled separately using the same linear mixed-effects structure as the primary analysis (scan condition, sonographer, and scan order as fixed effects; participant as a random intercept). Subscale comparisons are designated exploratory.

| Subscale | Manual mean | AI mean | Mean difference [95% CI] | *P* |
|---|---|---|---|---|
| Mental demand | 48.0 | 41.6 | −6.3 [−11.9, −0.8] | 0.03 |
| Physical demand | 42.2 | 38.6 | −3.6 [−9.5, +2.3] | 0.24 |
| Temporal demand | 33.8 | 34.8 | +1.1 [−5.4, +7.6] | 0.74 |
| Performance[1] | 45.2 | 42.3 | −2.9 [−10.4, +4.6] | 0.46 |
| Effort | 48.2 | 41.2 | −7.0 [−13.2, −0.7] | 0.04 |
| Frustration | 42.2 | 41.5 | −0.7 [−7.6, +6.2] | 0.84 |

*[1] Performance is scored such that lower values indicate higher self-rated performance; a negative AI-vs-manual difference therefore indicates better self-rated performance under AI assistance.*

**Table S8 — AI measurement generation and operator action**

Each of the six AI-supported measurements was attempted once in each of the 32 AI-assisted examinations, giving 192 measurement opportunities. For each, the table reports whether the AI generated a value and what the operator then did. Modification frequencies in Figure 5 are calculated against the AI generated column, since a measurement the AI never produced could not be modified.

| Measurement | AI generated | Modified | Measured manually after non-generation | Not obtained |
|---|---|---|---|---|
| Liver span | 31 | 18 | 0 | 1 |
| Gallbladder wall | 30 | 11 | 2 | 0 |
| Common bile duct diameter | 29 | 7 | 2 | 1 |
| Right kidney length | 30 | 5 | 1 | 1 |
| Left kidney length | 32 | 6 | 0 | 0 |
| Spleen length | 32 | 1 | 0 | 0 |
| **Total** | **184** | **48** | **5** | **3** |

*Modified = operator changed an AI-generated value. Measured manually after non-generation = the AI produced no value and the operator measured the structure. Not obtained = no measurement was recorded; one was a documented operator omission and two were not explained in the scan record. Rows do not sum to 32: examinations in which the AI-generated value was accepted unchanged are not shown.*

### Table S9 — AI-assigned and sonographer-amended anatomical label pairs

The 33 annotation modifications recorded during AI-assisted scanning, tabulated by AI-assigned label and sonographer-amended label. Pairs are grouped by anatomical region to indicate the structure of the modifications observed. Modifications were concentrated among anatomically adjacent, sonographically confusable structures, predominantly within the epigastric vascular and ductal region (17 of 33 modifications).

| Anatomical region | AI label | Sonographer-amended label | n |
|---|---|---|---|
| Epigastric vascular and ductal structures (n = 17) | Pancreas | IVC | 4 |
| | Left liver (long) | Aorta | 3 |
| | IVC | CBD | 2 |
| | IVC | Left liver (long) | 1 |
| | IVC | Right liver (long) | 1 |
| | IVC | Right liver (trans) | 1 |
| | CBD | Left liver (trans) | 1 |
| | CBD | Pancreas | 1 |
| | CBD | Gallbladder (long) | 1 |
| | Pancreas | CBD | 1 |
| | Aorta | Pancreas | 1 |
| Hepatic–renal and right upper quadrant (n = 5) | Right liver (long) | Right kidney (long) | 2 |
| | Right liver (long) | CBD | 1 |
| | Right liver (long) | Right liver (trans) | 1 |
| | Right liver (trans) | Right kidney (trans) | 1 |
| Renal laterality and views (n = 4) | Right kidney (long) | Left kidney (long) | 3 |
| | Left kidney (trans) | Right kidney (trans) | 1 |
| Gallbladder (n = 6) | Gallbladder (long) | Gallbladder (trans) | 1 |
| | Gallbladder (trans) | Gallbladder (long) | 1 |
| | Gallbladder (long) | Left kidney (long) | 1 |

| Anatomical region | AI label | Sonographer-amended label | n |
|---|---|---|---|
| | Gallbladder (trans) | Left kidney (long) | 1 |
| | Gallbladder (long) | Right liver (long) | 1 |
| | Gallbladder (trans) | Right liver (long) | 1 |
| Splenic (n = 1) | Spleen | Left kidney (trans) | 1 |
| **Total** | | | **33** |

### Supplementary Figures

#### Figure S1 — Hand-tracking system

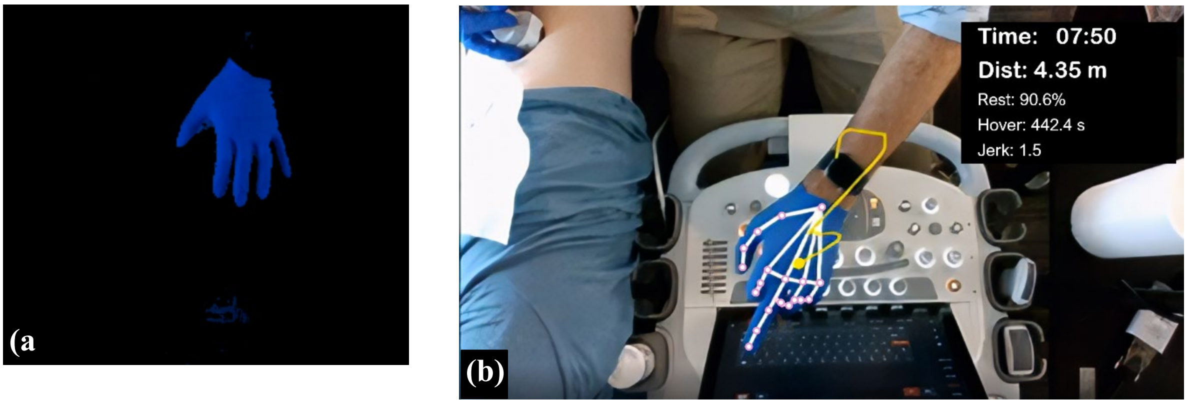


**Figure S1** — Hand-tracking system for quantifying hand–console interaction during ultrasound scanning. (a) Three-dimensional hand masking within the predefined workspace above the ultrasound console (blue overlay). (b) Centroid-based hand motion tracking, showing overlaid landmarks (pink points connected by white lines) and trajectory (yellow path). Quantitative metrics, including scan duration, cumulative hand travel distance, hover time, and average jerk, are displayed in real time.

**Figure S2 — Weighted NASA-TLX against scan duration**

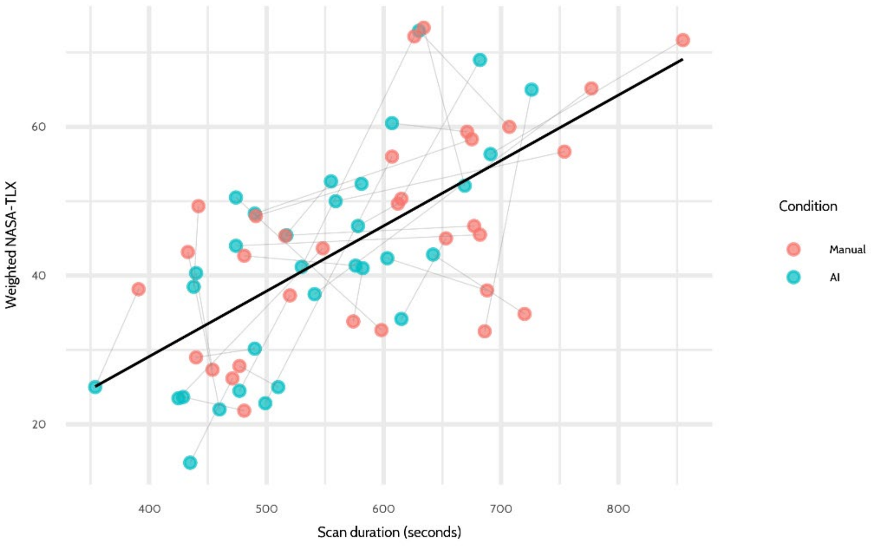


**Figure S2** — Weighted NASA-TLX against scan duration across all 64 examinations. Each dot is one scan, coloured by condition; grey lines connect the two scans for each participant. The black line shows the overall regression. Scan duration was associated with workload (β = 0.089 NASA-TLX points per second; 95% CI 0.060–0.118; $P < 0.001$) with no significant condition effect after adjustment for duration

**References**


1. Lin S-Z. HTS: Hand Tracking System. GitHub. https://github.com/LolMaple/HTS
2. Zhang F, Bazarevsky V, Vakunov A, et al. MediaPipe Hands: On-device Real-time Hand Tracking. 2020;https://doi.org/10.48550/arXiv.2006.10214.
3. Casiez G, Roussel N, Vogel D. 1 € filter: a simple speed-based low-pass filter for noisy input in interactive systems. presented at: Proceedings of the SIGCHI Conference on Human Factors in Computing Systems; 2012; Austin, Texas, USA. https://doi.org/10.1145/2207676.2208639
